# Learning to Personalize Medicine from Aggregate Data

**DOI:** 10.1101/2020.07.07.20148205

**Authors:** Rich Colbaugh, Kristin Glass

## Abstract

There is great interest in *personalized medicine*, in which treatment is tailored to the individual characteristics of patients. Achieving the objectives of precision healthcare will require clinically-grounded, evidence-based approaches, which in turn demands rigorous, scalable predictive analytics. Standard strategies for deriving prediction models for medicine involve acquiring ‘training’ data for large numbers of patients, labeling each patient according to the outcome of interest, and then using the labeled examples to learn to predict the outcome for new patients. Unfortunately, labeling individuals is time-consuming and expertise-intensive in medical applications and thus represents a major impediment to practical personalized medicine. We overcome this obstacle with a novel machine learning algorithm that enables *individual-level* prediction models to be induced from *aggregate-level* labeled data, which is readily-available in many health domains. The utility of the proposed learning methodology is demonstrated by: i.) leveraging US county-level mental health statistics to create a screening tool which detects individuals suffering from depression based upon their Twitter activity; ii.) designing a decision-support system that exploits aggregate clinical trials data on multiple sclerosis (MS) treatment to predict which therapy would work best for the presenting patient; iii.) employing group-level clinical trials data to induce a model able to find those MS patients likely to be helped by an experimental therapy.

## 1. Introduction

There is great interest in *personalized medicine*, in which treatment is tailored to the individual characteristics of patients [1-6]. This attention is motivated by a large array of expected benefits. For example, it is hypothesized that: i.) a particular drug will be most helpful if delivered to ‘the right patient at the right time’; ii.) the recent availability of population-scale data captured by electronic health records (EHRs), wearable sensors, and social media logs will allow the health of individuals to be monitored and even predicted. Achieving the goals of personalized medicine will require clinically-grounded, evidence-based solutions, which in turn implies that rigorous, scalable predictive analytics must play a central role in the path forward [3-6].

It is recognized that data-driven predictive modeling, especially *supervised machine learning*, is the preferred strategy when analyzing complex, uncertain systems (e.g. in terms of accuracy, reliability, and scalability) [7,8]. Given a phenomenon of interest, standard supervised learning begins with collecting and labeling example instances according to the outcome to be predicted (e.g. annotating patients as responders/non-responders for a drug). These labeled examples are then used to ‘train’ a model to predict the labels of new instances. Unfortunately, learning good models usually demands large numbers of labeled examples, and labeling data is time-consuming and expertise-intensive in medical applications. Indeed, the need to label individual patients represents a significant impediment to practical personalized medicine.

While *individual-level* (IL) patient labels are difficult to obtain, abundant *aggregate-level* labeled data already exists in many health domains. Observe, for example, that attempting to use the information contained in an EHR database to construct and publish a ‘disease profile’ for each patient in some population poses modeling challenges (e.g. mitigating EHR coding errors [9,10]) and raises ethical concerns (e.g. regarding patient privacy). In contrast, aggregate prevalence data is readily-available for many disorders (e.g. each year the US publishes such disease-prevalence statistics at the county-level [11]).

Recognizing the potential for aggregate data to facilitate development of evidence-based personalized medicine, we have derived a new approach to supervised learning which enables IL-prediction models to be induced from aggregate-level labels. That is, the proposed algorithm needs only group-level data for an outcome of interest (prevalence of breast cancer in US counties) to generate a model that predicts the outcome for a specific patient (it is likely Mary has breast cancer). Remarkably, we show through real-world case studies that the new learning scheme – which uses *no* IL labels for training – induces models with predictive performance that matches or exceeds state-of-the-art models trained on hundreds or thousands of labeled individuals. (The article [12] reports some of our early work on this idea.)

This paper describes how to leverage data labeled at the aggregate-level to enable delivery of effective personalized medicine and illustrates the method with experiments involving individualized disease-screening and precision therapy-selection. More explicitly, we

- offer an algorithm which uses the average rate at which a cohort experiences a health outcome to learn models capable of accurately predicting whether a given patient will experience that outcome;
- employ US county-level mental health statistics to learn a depression-screening tool that reliably identifies individuals suffering from depression [13,14] based upon their Twitter activity;
- design decision-support systems which exploit aggregate clinical trials data on multiple sclerosis (MS) drugs to: i.) predict which therapy would work best for a particular patient [15,16]; ii.) find probable responders to a candidate experimental therapy [17,18].

To gain intuition regarding the task of learning patient-level prediction models from aggregate data, we now provide a simple, informal comparison of the basic concepts behind standard supervised learning and our new learning technique. The cartoon in Figure 1 depicts the supervised learning process: given training examples, some labeled ‘red’ or ‘blue’ and some unlabeled (gray), the learner produces a model/decision boundary which predicts the classes of the unlabeled instances. With the new learning methodology, sketched in Figure 2, it is assumed no individual labels are known (all instances are gray). Instead, the only available evidence concerning class-membership of instances is for groups or ‘bags’ of individuals. Concretely, learning is guided by knowledge of the *distribution* of labels in some bags – here, the fractions of red and blue instances in the two leftmost bags (denoted by bars). Although learning is based upon aggregate-level label information, we seek a model able to make IL-level predictions (Figure 2, at right).

**Figure 1.**
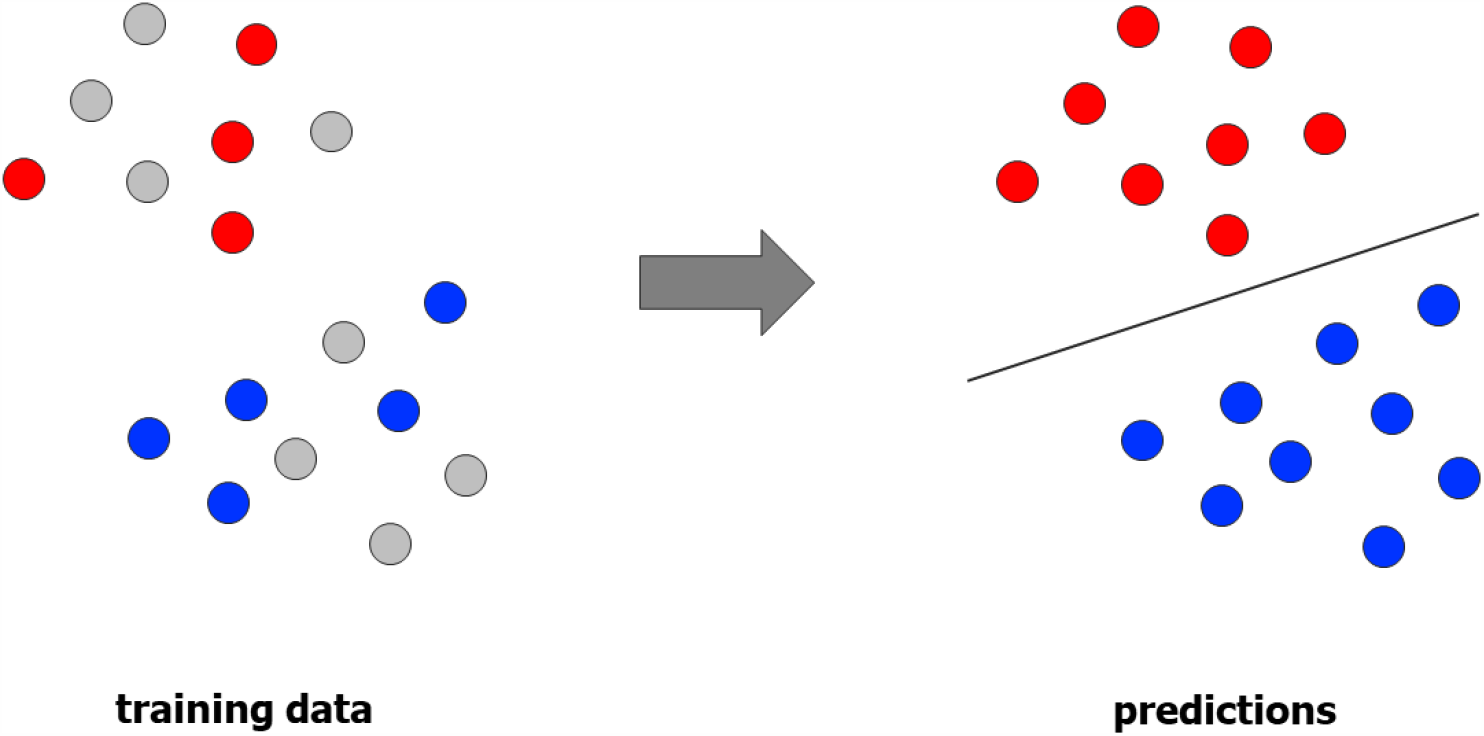
Standard supervised learning: IL prediction models are induced from IL labels.

**Figure 2.**
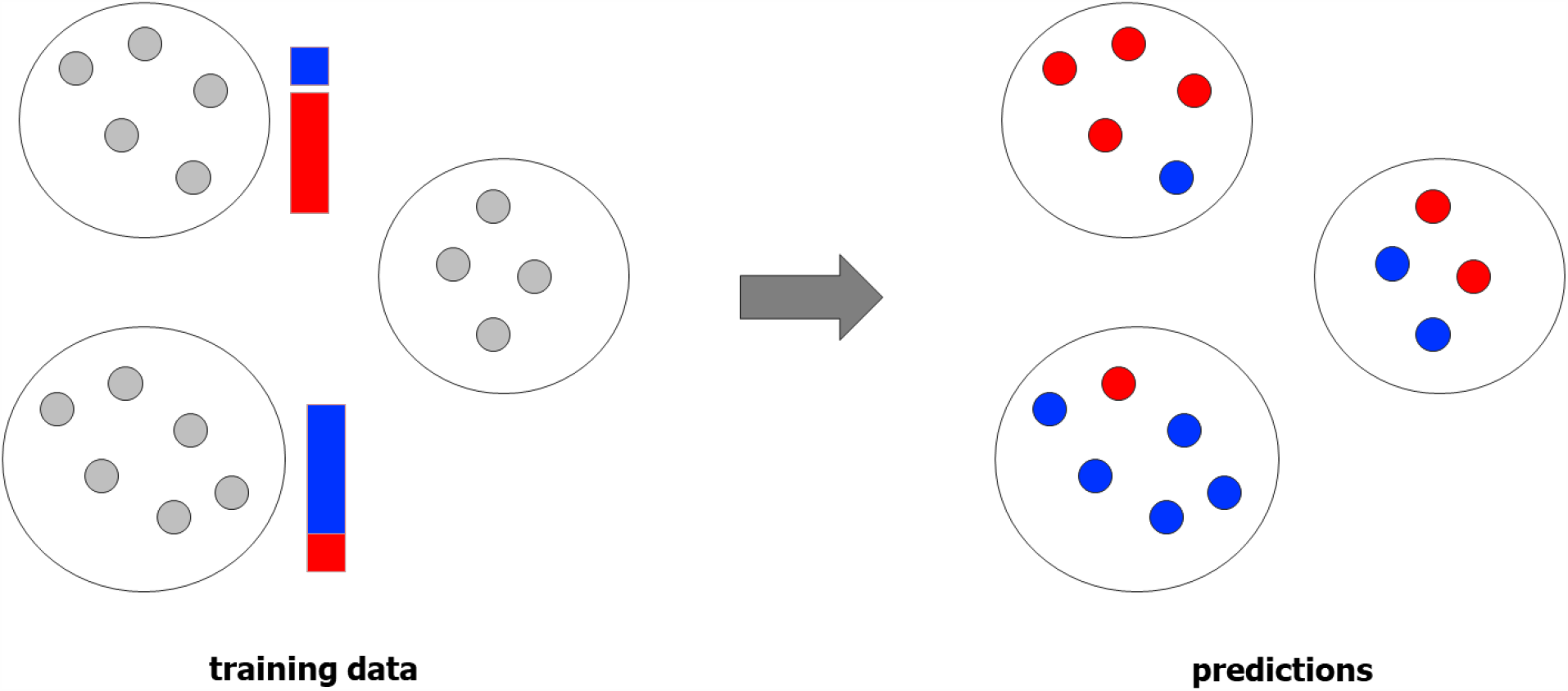
New learning method: IL prediction models are learned from aggregate labels.

The remainder of the paper is organized as follows. Section 2 presents the proposed approach to learning individual level models from aggregate data, and contains subsections on problem formulation, predictive modeling, and an introductory case study. Experiments validating the performance and utility of the new learning algorithm for personalized medicine are described in Sections 3-5 and include precision-screening for depression using Twitter activity (Section 3), individualized therapy selection for MS patients (Section 4), and responder/non-responder prediction for an experimental, as-yet-unapproved MS treatment (Section 5). Section 6 summarizes the paper and suggests directions for future work.

## 2. Learning Individual-Level Models from Aggregate Data

Constructing models to predict individual-level (IL) behavior via standard machine learning or bioinformatics techniques requires that large numbers of IL training examples be labeled in accordance with the outcome to be predicted [7,8,10]. In medical applications, labeling the outcomes of individuals (e.g. whether a patient has metastatic lung cancer) is time-consuming, demands domain expertise, and can give rise to privacy concerns. Consequently, obtaining IL labels represents a significant obstacle to deriving the data-driven models needed for personalized medicine. To overcome this obstacle, we have developed a new machine learning methodology which enables accurate IL prediction models to be learned from aggregate labels, that is, labels corresponding to group rather than individual outcomes (metastatic lung cancer rates for US states). This section of the paper presents the new learning process.

### A. Problem Formulation

We begin by defining the problem of interest. Given *aggregate* information about the (hidden) labels of individuals, the goal is to learn a model which accurately predicts *individual-level* labels. For simplicity it is supposed the prediction task is binary classification (e.g. deciding whether or not a patient is infected with a virus), but the analysis can be extended to multi-class classification and regression problems in a straightforward manner [12].

To state the problem quantitatively we introduce two data models, corresponding to the two levels of analysis:

- *instance-level:* the patient cohort of interest is modeled as D_I_ = {(**x**_1_, y_1_), …, (**x**_n_, y_n_)}, where **x** = [x_1_, …, x_d_]^T^ is the patient feature vector and, importantly, ground-truth labels y∈{−1,+1} are unknown; explicitly, the patient-level information available to the learner is *unlabeled IL data* D_IU_ = {**x**_1_, …, **x**_n_};
- *aggregate-level:* instances occur in ‘bags’ {**x**_i_ | i∈B_j_}, j∈{1, …, m}; each bag B_j_ is modeled by the mean **z**_j_ of instance feature vectors in that bag and is labeled with the bag’s fraction of positive instances f_j_ (assumed known), yielding the *labeled aggregate data* D_A_ = {(**z**_1_, f_1_), …, (**z**_m_, f_m_)}.

The problem is then: given labeled aggregate data D_A_ and unlabeled IL data D_IU_, predict ∀i the probability p_i_ that instance **x**_i_∈D_I_ has label y_i_ = +1.

### B. Learning Method

The proposed approach to learning IL predictions from aggregate labels (LIPAL) consists of three steps: representation learning, aggregate-based prediction, and clustering-informed prediction refinement (see Figure 3). We now describe each step in the procedure and then specify the complete LIPAL algorithm.

**Figure 3.**
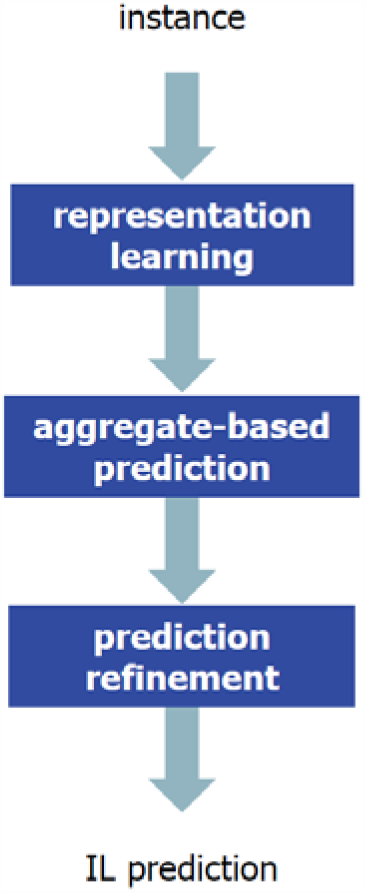
Schematic of new method for making IL predictions using models learned from aggregate data.

#### Representation learning

The first step in the prediction process is to transform the original, ‘raw’ instance representation **x**°∈ℜ^do^ to the form **x** = [x_1_, …, x_d_]^T^ used in data model D_I_, where feature vector **x** better captures the underlying structure of the data [7]. We employ two techniques to learn the transformation **x**° → **x**: Latent Dirichlet Allocation (LDA) [19] and deep learning with stacked autoencoders (SAE) [20]. In each case, it is assumed the data distribution from which D_I_ is sampled concentrates near a low-dimensional manifold, and the objective is to identify ‘latent’ variables that characterize this manifold.

For example, analysis of scientific literature often focuses on the content of papers, and it is common to model each paper as a bag of words **x**°∈ℜ^| V|^, where the entries of **x**° are the frequencies with which words in vocabulary V appear in the paper. The latent features identified by LDA then have a natural interpretation as topics being discussed in a corpus of papers (see Section 2.C for an application to social media).

Alternatively, with SAE the goal is to learn a parsimonious encoding of **x**°, **x** = f_θ_(**x**°), which permits accurate decoding **x**_d_ = g_θ_(**x**) ≈ **x**°, where f_θ_ and g_θ_ are encoding and decoding functions, respectively, and θ is the vector of parameters to be learned [20]. Note that both LDA and SAE learn feature representations in an unsupervised fashion, so each can be implemented directly with data D_IU_ (recall instance-level labels y are unknown, so learning must be unsupervised). Because **x** encodes the manifold structure of the data, these new features are typically more informative for predictive modeling than raw features **x**° [7,8,19-23].

#### Aggregate-based prediction

The second step in the prediction process is to leverage the ‘light’ supervision provided by labeled aggregate data D_A_ to induce a model which enables preliminary IL predictions to be made. The prediction model is obtained through a two-stage learning procedure:

- aggregate-level regression: given labeled aggregate data D_A_ = {(**z**_1_, f_1_), …, (**z**_m_, f_m_)}, learn an ensemble of decision trees [7] regression model **f**_r_: Z → F that accurately predicts the fraction of positive instances f_*_ in a new (unseen) bag B_*_;
- instance-level prediction: apply the aggregate-level model **f**_r_ to the instances {**x**_1_, …, **x**_n_} in D_IU_ to form predictions **p**_0_ = [p_01_, …, p_0n_]^T^, where p_0i_ is the predicted probability that instance **x**_i_ has label y_i_ = +1.

Because predictions **p**_0_ are derived using a model learned on the mean behaviors (**z**_j_, f_j_) of bags of instances, they are on average good estimates for the probabilities that instances **x**_i_ have positive label [22]. However, these predictions may have large variance (e.g. if bags are few in number or lack diversity), motivating a prediction-refinement step.

#### Prediction refinement

The third step in the prediction process involves refining preliminary predictions **p**_0_ to final predictions **p** = [p_1_, …, p_n_]^T^, where p_i_ is the predicted probability **x**_i_ has label y_i_ = +1. The idea behind the refinement process is that if instances **x**_k_, **x**_l_ are contained in the same bag and are ‘similar’, then they should possess similar labels. This reasoning suggests that unlabeled IL data, if informative concerning instance similarity, may be helpful in improving predictions **p**_0_.

Let L_j_=I−C_j_, where I is the identity matrix and C_j_ is a similarity matrix computed via ensemble clustering [7,12] on bag B_j_. Specifically, we construct matrix C_j_ so that its (k,l) entry is equal to the number of times B_j_’s instances **x**_k_, **x**_l_ are assigned to the same cluster by members of the ensemble. Final predictions **p** are formed by optimally balancing the goals of maintaining agreement with preliminary predictions **p**_0_ and achieving within-cluster label similarity:

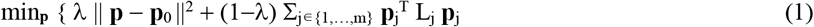

subject to the constraint p_i_∈[0,1] ∀i, where **p**_j_ is the subset of predictions **p** corresponding to bag B_j_ and λ∈[0,1] reflects the relative expected predictive value of aggregate-level label data and instance-level clustering.

It is seen that the predictions **p** generated by minimizing (1) incorporate information from three sources: unsupervised IL representation learning (**x**° → **x**), supervised aggregate-level learning (via **p**_0_), and unsupervised IL clustering (through the L_j_). The optimization (1) can be accomplished, independently for each bag, by iterating the following formula over index i until convergence (which is guaranteed [21]):

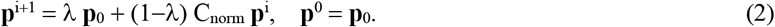

In (2), C_norm_ is obtained by normalizing C to a symmetric probability matrix [21], and bag index j is suppressed for clarity. This solution is efficient to compute, allowing large-scale problems to be investigated.

We summarize the discussion by sketching the proposed algorithm for making IL predictions.

#### Algorithm LIPAL

1. Learn a representation **x** for feature vector **x**° which captures the underlying structure of IL data D_IU_ (e.g. using LDA [19] or SAE [20]).
2. Use aggregate-level labeled data D_A_ to learn an ensemble of decision trees regression model **f**_r_ that accurately predicts the fraction of positive instances f_*_ in a new bag B_*_.
3. Compute preliminary predictions **p**_0_ for all the instances in D_IU_ using **f**_r_.
4. Perform prediction refinement **p**_0_ → **p** by optimizing (1) using iteration (2).

The performance of Algorithm LIPAL is now briefly illustrated with a case study on income prediction (see [22] for a more detailed account of the study).

### C. Case Study: Income Prediction

Consider the task of using geographically-aggregated income data and IL social media posts to learn a model which detects low-income individuals based only on their Twitter activity. The data employed to learn the prediction model and evaluate its performance consists of geo-tagged Twitter posts and income levels for 5191 UK/US residents, 500 with low income and 4691 with typical earnings, collected during 2014 [24]. We test our method by aggregating this ‘UK/US cohort’ into 20 city-level bags with the scheme reported in [25], using this aggregate data to learn a model for detecting low-income users from their Twitter posts via Algorithm LIPAL, and then checking these predictions against the IL income labels from [24]. It is emphasized that IL labels are kept hidden during learning and used only for model evaluation.

Algorithm LIPAL is implemented to solve this prediction problem in the following way:

- representation learning: use LDA [19] to learn the 200 primary topics being discussed by the UK/US cohort on Twitter; the extent to which user i posts about each of the topics, together with the user’s age/gender/ethnicity, estimated from users’ posts as in [22], defines the d = 203 elements of feature vector **x**_i_;
- aggregate-based prediction: learn model **f**_r_ (ensemble of 1000 regression trees [7]) on labeled aggregate data D_A_ where, for each bag B_j_, **z**_j_ is the mean Twitter-based feature vector and f_j_ is the fraction of low-income individuals in that bag; apply **f**_r_ to users {**x**_1_, …, **x**_5191_} in D_IU_ to form preliminary IL predictions **p**_0_;
- prediction refinement: map **p**_0_ → **p** by optimizing (1) using (2) with λ = 0.3 (λ is set by approximate tuning on a small held-out validation set).

For comparison purposes, three other prediction models are examined: ‘demographics’ and ‘topics’, induced by running Algorithm LIPAL on just demographic-features or topic-features, respectively, and ‘instance-labeling’, a state-of-the-art random forest (RF) classifier [7] trained on 3000 IL-labeled users (these users are modeled with the same 203 features used by Algorithm LIPAL). Predictive performance is measured with class-average (CA) accuracy, estimated through cross-validation. (Recall CA accuracy averages sensitivity and specificity and remains informative even when the data is class-imbalanced [7].)

The results obtained in this case study are displayed in Figure 4. It can be seen that Algorithm LIPAL, which has access to *no* IL labels, achieves CA accuracy (90.1%) comparable to a state-of-the-art classifier trained with 3000 IL-labeled users (CA accuracy = 90.5%). (Note that 3000 labeled training instances are needed for the RF classifier to attain a performance-level similar to Algorithm LIPAL; using fewer IL labels yields lower accuracy.)

**Figure 4.**
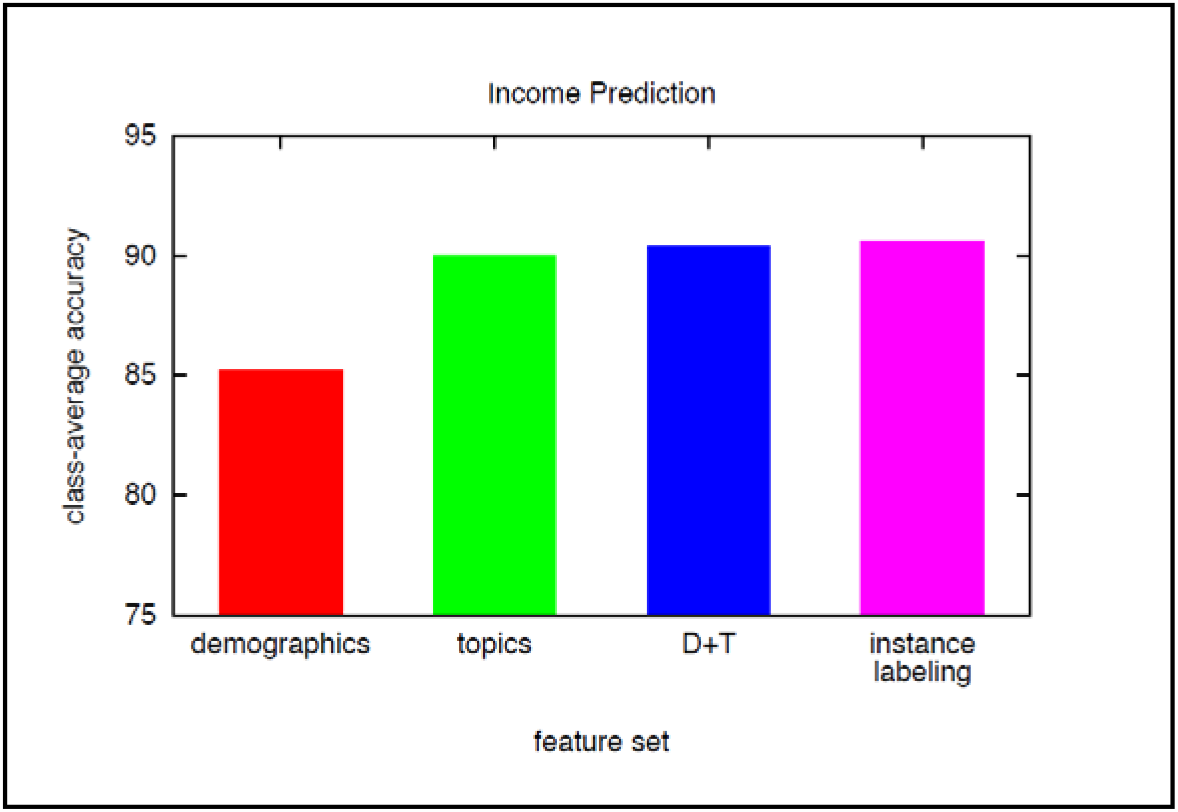
Income prediction. Plot shows accuracy of Algorithm LIPAL using demographic features (red), topic features (green), and both demographics and topics (blue); also shown is accuracy of an RF classifier trained on 3000 individually-labeled users (magenta).

## 3. Experiment One: Personalized Depression Screening

We now describe an empirical investigation in which Algorithm LIPAL is used to predict whether a given individual has depression based upon that person’s Twitter activity. Depression is a leading cause of disability worldwide, estimated to affect more than 350M people [26]. Affected patients can experience intermittent symptoms, social stigma, and impaired decision-making, which can in turn impede diagnosis and inhibit care-seeking. As a consequence, there is great interest in developing *personalized, passively-deployable* depression-screening systems, based upon analysis of social media data, in order to facilitate earlier detection and more effective treatment [13,14,27,28].

A central challenge in building such screening tools is assembling IL-labeled training data – users for whom social media data has been collected and depression status has been determined [14,26-28]. We therefore examine the utility of exploiting *aggregate* mental health data with Algorithm LIPAL to learn to detect depressed individuals from their social media activity.

### A. Setup

The goal of these experiments is to learn models which can accurately predict whether an individual has depression based upon the person’s Twitter activity. The three sources of data are used in this experiment are listed below.

- Individual-level Twitter activity dataset D_I-Twitter_ = {**x**_1_, …, **x**_840_}, collected for 840 users during 2013 via Amazon Mechanical Turk [29] following the protocol in [14]; feature vector **x** = [x_1_, …, x_189_]^T^ encodes the 188 user-attributes identified in [14] plus the user’s US state of residence (inferred with the technique in [25]).
- State-level depression dataset D_A-depression_ = {(**z**_1_, f_1_), …, (**z**_20_, f_20_)}, downloaded for the 20 states in which the users contained in D_I-Twitter_ reside [30]; f_j_ is the depression rate for state j in 2013 and **z**_j_ is the mean Twitter feature vector for the users in D_I-Twitter_ who live in that state.
- Individual-level depression-label dataset D_I-PHQ_ = {(**x**_1_, y_1_), …, (**x**_840_, y_840_)}, where label y_i_∈{−1,+1} reflects the result of a PHQ-9 assessment administered to user **x**_i_ when compiling **x**_i_’s Twitter data [31]; 306 participants are labeled ‘depressed’ (PHQ-9 score ≥ 10) and 534 are labeled ‘not depressed’ (PHQ-9 < 10) [26] (these labels are used *only* for model evaluation).

These datasets illustrate the challenges faced when analyzing social media for personalized medicine [14,27,28]. For instance, it is typically straightforward to acquire unlabeled user-activity logs (e.g. Twitter posts) and aggregate statistics on labels of interest (depression rates in US states), but much more difficult to gather mental health diagnoses for individuals; as indicated above, the latter are needed to guide existing social media modeling schemes [e.g. 14].

The study procedure consists of two components: model learning and model evaluation. Crucially, IL labels are used only to assess the performance of prediction models and not during model induction. Thus, in the predictive modeling stage, a depression-detection model is generated by applying Algorithm LIPAL to the unlabeled Twitter activity dataset D_I-Twitter_ and aggregate-label/state-level depression dataset D_A-depression_. The resulting model is then evaluated by leveraging ground-truth IL-labeled dataset D_I-PHQ_ = {(**x**_1_, y_1_), …, (**x**_840_, y_840_)}.

In these experiments, Algorithm LIPAL is implemented in the three steps specified in Section 2.B and summarized below:

1. representation learning: we skip this step and instead adopt the feature vectors engineered in [14] to allow direct comparison with that work;
2. aggregate-based prediction: form IL predictions **p**_0_ for users in D_I-Twitter_ with the model **f**_r_ learned on state-level dataset D_A-depression_ (**f**_r_ is an ensemble of 1000 regression trees [7]);
3. prediction refinement: map **p**_0_ → **p** using iteration (2) (with λ = 0.3) and unlabeled data D_I-Twitter_.

### B. Results

Algorithm LIPAL is now applied to the task of predicting whether a given individual has depression based on their Twitter activity. For comparison purposes, three other prediction models are examined:

- ‘network’, constructed by running Algorithm LIPAL on just Twitter network features (e.g. number of followers and accounts followed, reciprocity, graph density/embeddedness/clustering coefficient – see [14] for a full list);
- ‘language’, built via Algorithm LIPAL with just Twitter post features (e.g. number of posts/replies, level of positive affect/negative affect/activation, use of first-person pronouns and swearing – see [14] for a full list);
- ‘horvitz’, a state-of-the-art depression-detection model [14] trained on 600 IL-labeled users (modeled with the same 189 features used by Algorithm LIPAL).

Predictive performance is measured with CA accuracy and area under ROC curve (AUC) estimated through cross-validation [7].

The results of this experiment are displayed in Figure 5. It is seen that Algorithm LIPAL, which has access to *no* IL labels, is able to accurately predict depression status of users (magenta, CA accuracy = 0.88 and AUC = 0.94). Using only Twitter network features (blue) or post features (green) also permits useful prediction. All these models are substantially more accurate than a state-of-the-art depression classifier [14] trained on hundreds of IL labels (red).

**Figure 5.**
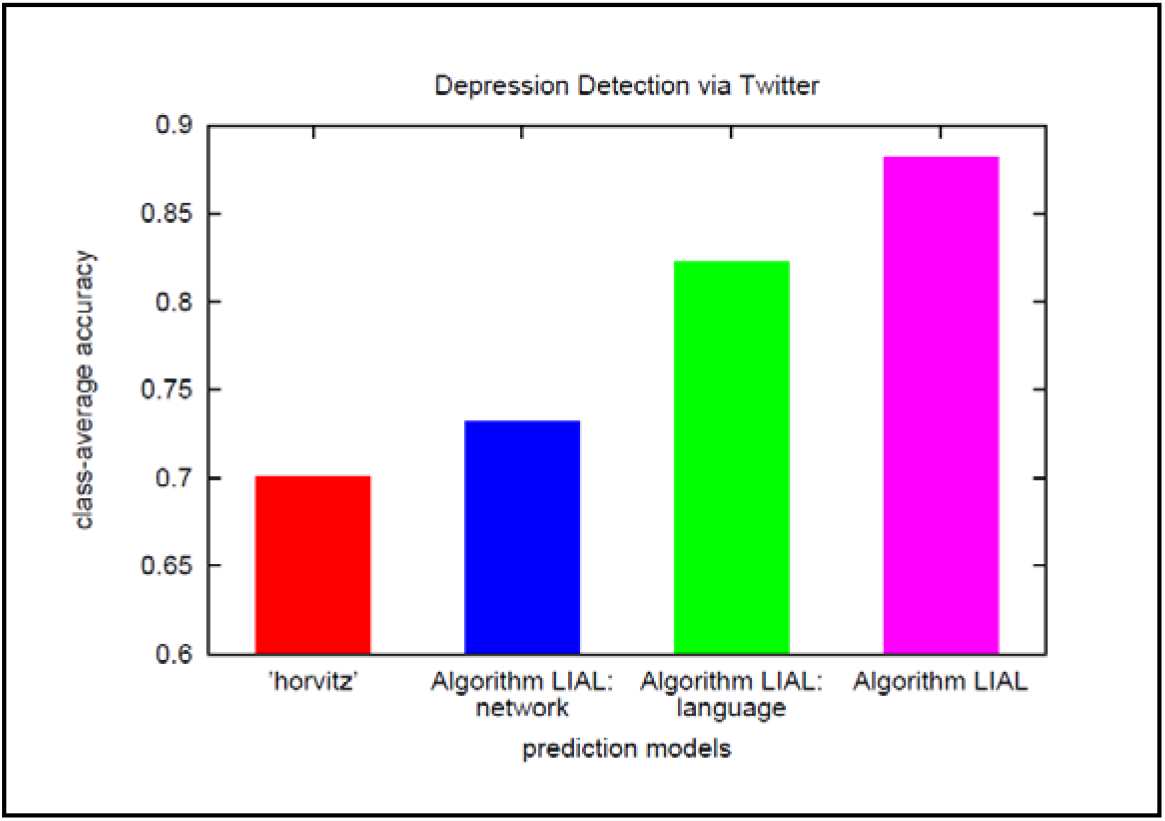
Depression prediction. Plot shows accuracy of Algorithm LIPAL using Twitter network features (blue), language features (green), and both network and language (magenta); also shown is the accuracy of classifier [14] trained on 600 individually-labeled users (red).

Knowing which features captured by Twitter are predictive of depression is of value in various clinical applications. We assessed feature predictive power using both forward- and backward-stepwise analyses [7], and now highlight a few findings. Especially predictive language features include: i.) increased use of negative affect, first-person pronouns, and swearing; ii.) decreased emotional activation. Among the network features with predictive power are: i.) increased ego-network clustering coefficient and embeddedness; ii.) decreased ego-network size and overall social engagement (see [14] for definitions of all terms).

## 4. Experiment Two: Personalized MS Therapy Selection

This section presents experiments in which Algorithm LIPAL is used to predict which of a set of MS therapies will deliver the optimal outcome for a particular individual. MS is a disorder of the central nervous system with substantial individual and societal costs. As there is currently no cure, care emphasizes timely initiation of suitable therapy to slow or perhaps even halt disability progression. However, choosing appropriate intervention is difficult because treatment response varies widely across patients [15,16].

While *evidence-based personalized therapy* is therefore desirable, realizing this objective demands models able to accurately predict individual responses to approved disease-modifying therapies (DMTs). Because of the complexity and heterogeneity of the MS phenotype, inducing these models via standard methods requires response data for large numbers of patient-DMT pairs and this is usually difficult to obtain [15,32,33]. Here we examine the possibility that already-collected clinical trials data – which is aggregated to protect privacy of participants and commercial interests of drug-makers – could be exploited with Algorithm LIPAL to produce models that predict individual-level response to MS DMTs.

### A. Setup

The goal of these experiments is to build models which would assist clinicians with the task of prescribing DMTs to accomplish one of two core MS objectives: i.) reduce risk of disability progression, or ii.) prevent conversion to the secondary progressive form of MS. More precisely, we wish leverage already-available, aggregate clinical trials data to derive models capable of predicting which of a set of candidate DMTs will be best for a given individual. Four relapsing-remitting MS drugs are investigated – interferon beta (IB), glatiramer acetate (GA), fingolimod (FIN), and natalizumab (NAT) [32,33]. This specification of therapy targets and candidate DMTs guided our selection of, and data extraction from, 11 relevant clinical trials (clinicaltrials.gov):

- most pertinent: NCT01633112 [1064 patients], NCT00340834 [1292 patients], NCT02342704 [108 patients], NCT01333501 [151 patients], NCT01534182 [298 patients], NCT00078338 [764 patients], and NCT01058005 [75 patients];
- supporting trials: NCT00605215 [881 patients], NCT00451451 [1417 patients], NCT00027300 [900 patients], and NCT01569451 [53 patients].

For each of the two treatment endpoints of interest, three categories of data are modeled as outlined below.

- Individual-level MS patient dataset D_I-MSBase_ = {**x**_1_, …, **x**_8513_}, collected for 8513 individuals in the MSBase registry [34], where each patient vector **x**∈ℜ^10^ is the SAE-transformed version of raw feature vector **x**° = [x°_1_, …, x°_19_]^T^ encoding the 19 demographic/clinical/paraclinical attributes suggested in [34,15].
- Four aggregate treatment response datasets D_A-IB_, D_A-GA_, D_A-FIN_, D_A-NAT_. Each dataset is of the form D_A-*_ = {(**z**_1_, f_1_), …, (**z**_K_, f_K_)} and contains aggregate treatment outcomes for patient-bags derived from the 11 clinical trials above (K = 23, 21, 20, 16 for IB, GA, FIN, NAT, respectively); f_j_ is the trial-reported responder rate for bag j (for IB, GA, FIN, or NAT) and **z**_j_ is the mean feature vector for patients in that bag.
- Individual-level treatment-response dataset D_I-treat_ = {(**x**_1_, y_1_), …, (**x**_8513_, y_8513_)}, where label y_i_∈{0,1}^4^ records the best therapy for patient **x**_i_ among the candidate drugs IB, GA, FIN, NAT (inferred from MSBase [33,15]); for example, y_9_ = [0,0,1,0] implies patient 9 responds best to FIN (these labels are used *only* to evaluate predictions of the learned models).

These datasets exemplify the challenges of analyzing clinical trials and disease-registry data for personalized medicine [15,32-34]. For instance, while it can be straightforward to gather unlabeled patient-attribute data (e.g. from observational studies) and aggregate statistics on treatment-response (from trials), it is usually more difficult to obtain response-labels for individuals; as mentioned above, the latter serve as inputs to standard supervised learning algorithms [7,8].

For each treatment goal – reduce risk of disability progression or prevent conversion to secondary progressive MS – the study procedure consists of three components: model learning, DMT optimization, and performance evaluation. Crucially, IL labels are used only to assess the prediction outcomes and not during model induction. Thus, in model-learning, four response-prediction models are generated, one for each DMT, by applying Algorithm LIPAL to unlabeled MS patient dataset D_I-MSBase_ and the relevant aggregate-response dataset D_A-*_. The models are then used to select, for each patient, the DMT predicted to work best. Finally, DMT optimization quality is assessed by comparing the predictions against ground-truth IL-labeled dataset D_I-treat_ = {(**x**_1_, y_1_), …, (**x**_8513_, y_8513_)}.

In these experiments, Algorithm LIPAL is implemented in the three steps specified in Section 2.B and summarized below:

1. representation learning: map **x**° → **x** with the SAE-based process proposed in [23] (see also [8,20] for background on SAEs);
2. aggregate-based prediction: form IL predictions **p**_0-IB_, **p**_0-GA_, **p**_0-FIN_, **p**_0-NAT_ for the patients in D_I-MSBase_ with ensemble regression models **f**_r-IB_, **f**_r-GA_, **f**_r-FIN_, **f**_r-NAT_ learned on aggregate-label datasets D_A-IB_, D_A-GA_, D_A-FIN_, D_A-NAT_ (each of the **f**_r-*_ is an ensemble of 1000 regression trees and missing feature values are imputed with a tree-based scheme [7]);
3. prediction refinement: map **p**_0-*_ → **p**_*_ using iteration (2) (with λ = 0.3) and unlabeled data D_I-MSBase_.

### B. Results

Algorithm LIPAL is first deployed for the task of predicting which DMT will optimally reduce the risk of MS disability progression for each of the 8513 patients in D_I-MSBase_. The results of the experiment are displayed in Figure 6. The left plot shows the predictions of Algorithm LIPAL and the right plot is the output of state-of-the-art model [15] trained on 7500 IL-labeled patients. Each plot is a two-dimensional feature-space projection with (predicted) optimal DMTs indicated by color: GA is cyan, FIN is green, and NAT is yellow. Notice that the predictions of Algorithm LIPAL, which needs *no* IL labels for training, are in close agreement with those of a high-quality model [15] trained on thousands of IL labeled patients.

**Figure 6.**
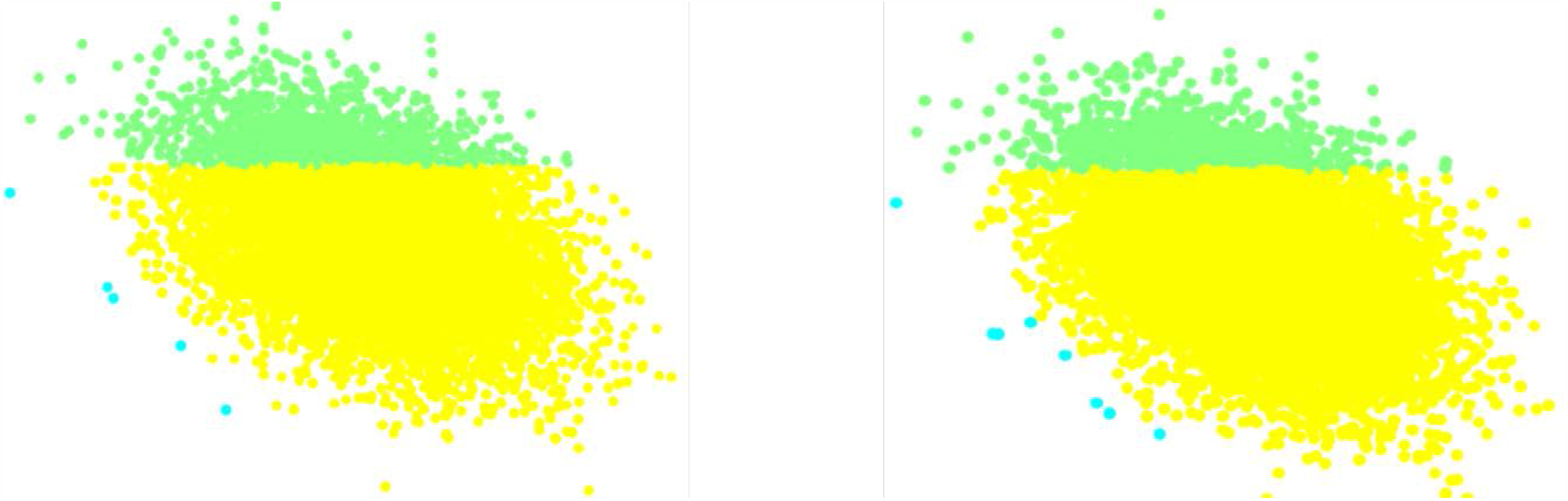
DMT selection to optimally-reduce risk of MS disability progression. Left plot shows predictions of Algorithm LIPAL and right plot is output of state-of-the-art model [15] trained on 7500 individually-labeled examples. In each case, plot is two-dimensional feature-space projection and optimal DMT for each patient is indicated by color: GA is cyan, FIN is green, and NAT is yellow.

Next we examine the ability of Algorithm LIPAL to predict which of the candidate DMTs will most effectively prevent conversion to secondary progressive MS for each of the 8513 patients in D_I-MSBase_. The findings of this experiment are displayed in Figure 7. The plot is a two-dimensional feature-space projection showing the optimal DMTs as predicted by Algorithm LIPAL (GA is cyan and NAT is yellow). Again the output of Algorithm LIPAL, trained without IL labels, is in close agreement with model [15] trained on thousands of IL labels (output for model [15] is indistinguishable for this experiment and thus is not shown).

**Figure 7.**
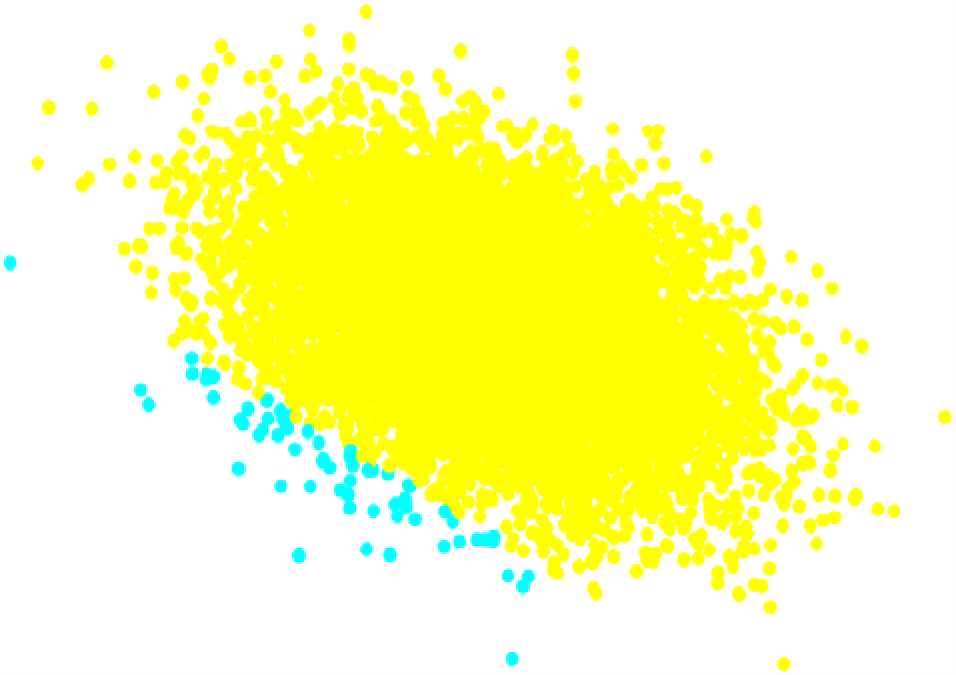
DMT selection to optimally-prevent conversion to secondary progressive MS. Plot is a two-dimensional feature-space projection of the predicted best therapy according to Algorithm LIPAL (GA is cyan and NAT is yellow).

Finally, an exploratory experiment was conducted to test the feasibility of applying Algorithm LIPAL to aggregate clinical trials data in order to forecast the number of disability-progression events each patient in D_I-MSBase_ will experience in the coming year. The results of this study are displayed in Figure 8. The plot depicts forecasting error rates for models obtained using Algorithm LIPAL for different DMTs (IB/green, GA/blue, FIN/magenta, NAT/cyan) as well as the error rate of a baseline model which simply predicts the training sample cohort mean (red, average error over all DMTs). Also plotted is error rate for the DMT mitoxantrone [33] (yellow); achieving good results for mitoxantrone suggests Algorithm LIPAL is generalizable to DMTs beyond those investigated here.

**Figure 8.**
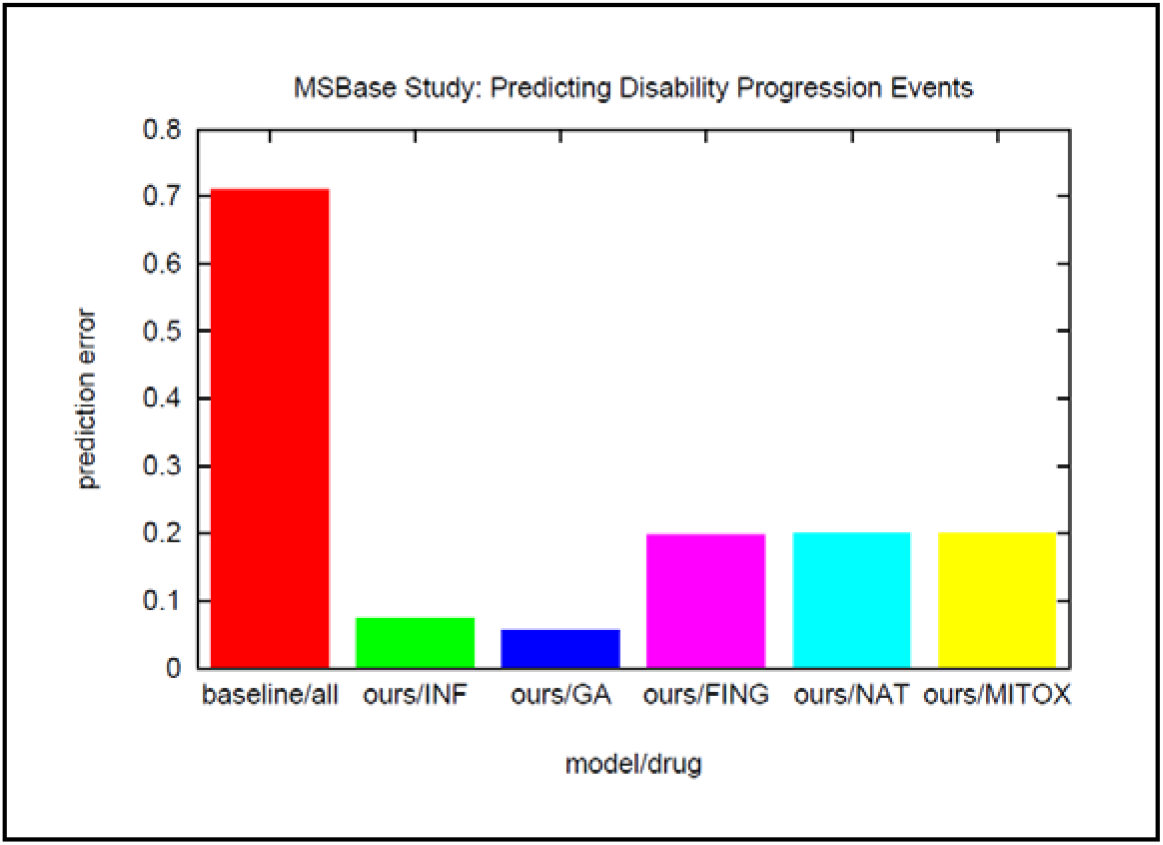
Forecasting number of disability-progression events in the coming year. Plot displays average prediction errors for patients in D_I-MSBase_ for baseline model (red bar, average error over all DMTs) and for Algorithm LIPAL (IB/green, GA/blue, FIN/magenta, NAT/cyan, and mitoxantrone/yellow).

Knowing which features captured in clinical trials data are predictive of DMT-efficacy is of value in several medical domains. We estimate feature predictive power using forward/backward-stepwise analyses [7] and now highlight a few findings. Intriguingly, features which predict therapy-effectiveness vary across the DMTs. For example, predictive features for utility of FIN and NAT include Expanded Disability Status Scale (EDSS) score, preceding year’s relapses, functional system score (FSS, e.g. pyramidal/cerebellar), and annual relapse rate. Features which are useful when assessing the value of treatment with IB and GA comprise EDSS, FSS (brainstem/cerebral), preceding year’s relapses, and number of relapses on treatment.

Interestingly, knowledge of features with predictive power can enable construction of prediction models which are simpler and more interpretable than those generated by Algorithm LIPAL [33]. To illustrate the basic idea, consider the problem of deciding which of the two drugs FIN or NAT would likely work better for reducing risk of disability progression for a given patient. Abstracting the model produced by Algorithm LIPAL using information about predictive features yields the prediction model summarized in the text box below (see [33,35] for details).

#### Simplified Therapy Selection Rule: FIN vs NAT

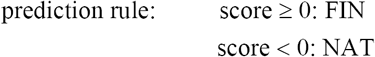

where

score = 15 * # relapses previous year − 5.8 * age − 5.7 * relapse-remit status − 5.4 * EDSS −

4.8 * FSS pyramidal − 4.1 * FSS cerebellar − 4.3 * FSS sphincteric − 5.1 * FSS ambulatory.

## 5. Experiment Three: MS Therapy Responder Detection

This section presents experiments in which Algorithm LIPAL is used to predict which patients, if any, would benefit from treatment with an experimental MS DMT. It is hypothesized that some DMTs which are unsuccessful in clinical trials may nevertheless represent effective MS treatments for certain subpopulations [3,17,18,32,33]. If true, taking advantage of this opportunity requires models capable of distinguishing responders from non-responders based upon patient attributes observable *before* commencing therapy.

The responder-prediction models required to exploit this potential opportunity in clinical settings must be: i.) learnable from response data which is already-available or easy to obtain [33]; ii.) validated with patient cohorts not ‘seen’ during model-training [3,33]. This experiment examines the possibility that combining Algorithm LIPAL with existing, aggregate clinical trials data can produce such models and thereby facilitate reliable identification of individuals likely be helped by a given MS DMT.

### A. Setup

It is sometimes the case that, although an experimental MS DMT fails to demonstrate efficacy for the operative clinical trial endpoint(s), there is reason to suspect the drug may benefit certain patient-subpopulations. This experiment investigates whether applying Algorithm LIPAL to already-collected aggregate clinical trials data generates models which enable detection of DMT-responders in a new (unseen) cohort. In this study the drug of interest is laquinimod [18,33] and the treatment goal is to reduce risk of disability progression.

Guided by this specification of DMT and therapy target, we chose four relevant clinical trials (clinicaltrials.gov) and one related scientific paper [18] and then extracted aggregated laquinimod-response data from these sources:

- trials: NCT00509145 (ALLEGRO) [1101 patients], NCT00605215 (BRAVO) [881 patients], NCT01707992 (CONCERTO) [1456 patients], NCT02284568 (ARPEGGIO) [374 patients];
- paper: Bovis, F et al., *BMC Medicine*, Vol. 17, 2019 [3438 patients] [18].

This data was processed to yield the three data models delineated below.

- Individual-level MS patient dataset D_I-CONCERTO_ = {**x**_1_, …, **x**_1456_} for 1456 patients, reverse-engineered from the CONCERTO clinical trial [NCT01707992], where each feature vector **x** = [x_1_, …, x_9_]^T^ encodes 9 demographic, clinical, and paraclinical patient attributes suggested in [18] (see also [33]).
- Aggregate laquinimod-responder dataset D_A-laquinimod_ = {(**z**_1_, f_1_), …, (**z**_12_, f_12_)} containing treatment outcome statistics for 12 bags of patients derived from the paper [18] and three clinical trials ALLEGRO [NCT00509145], BRAVO [NCT00605215], and ARPEGGIO [NCT02284568]; f_j_ is the trial-reported responder rate for bag j and **z**_j_ is the mean feature vector for patients in that bag.
- Individual-level laquinimod-response dataset D_I-Bovis_ = {(**x**_1_, y_1_), …, (**x**_1456_, y_1456_)}, where label y_i_∈{−1,+1} denotes responder/non-responder status for patient **x**_i_ in the CONCERTO trial (labels acquired from [18]); adopting disability progression-free survival as the metric, 441 patients are labeled ‘responders’ and 1015 patients are ‘non-responders’ (these labels are used *only* for model evaluation).

These datasets exemplify the challenges encountered when attempting to analyze clinical trials data for personalized medicine [17,18,32,33]. For instance, it is not hard to gather unlabeled patient-attribute data and aggregate statistics on treatment outcomes (e.g. responder rates in a clinical trial), but it is typically more difficult to obtain such labels for individuals.

Our study procedure consists of two components: model learning and model evaluation. Crucially, IL labels are used only to assess the performance of prediction models and not during model induction. Thus, in the modeling stage, a responder-prediction model is generated by applying Algorithm LIPAL to unlabeled MS patient dataset D_I-CONCERTO_ and aggregate-responder dataset D_A-laquinimod_. The predictions of the resulting model for D_I-CONCERTO_ participants are then assessed by comparing against ground-truth IL-labeled dataset D_I-Bovis_ = {(**x**_1_, y_1_), …, (**x**_1456_, y_1456_)}.

In the experiments, Algorithm LIPAL is executed in three steps as specified in Section 2.B and summarized below:

1. representation learning: skip this step and instead use feature definitions in [18] to allow direct comparison with that work;
2. aggregate-based prediction: form IL predictions **p**_0_ for patients in D_I-CONCERTO_ with the model **f**_r_ learned on aggregate labels D_A-laquinimod_ (**f**_r_ is an ensemble of 1000 regression trees and any missing feature values are imputed with a tree-based scheme [7]);
3. prediction refinement: map **p**_0_ → **p** using iteration (2) with λ = 0.3.

### B. Results

Algorithm LIPAL is now applied to the task of predicting which CONCERTO trial participants would be responders to laquinimod – that is, would have less disability progression with the drug than with a placebo – using the data collected during the earlier laquinimod trials ALLEGRO, BRAVO, and ARPEGGIO (data from the latter two trials was used to inform the grouping of patients into bags [33]). This problem formulation accommodates two important constraints associated with practical DMT responder-detection: i.) model-development is performed with data assembled *prior* to the target clinical trial [3,18,33]; ii.) the prediction model is learned from *aggregate-level* labels [33].

To place the experimental results in context, two views of data gathered in the ALLEGRO and CONCERTO trials are offered in Figures 9 and 10. (More details on the data can be found in [18] and clinicaltrials.gov.) Figure 9 depicts two-dimensional feature-space projections of ground-truth responders (yellow) and non-responders (blue) for the ALLEGRO trial (left) and CONCERTO trial (right) [18] (recall these IL labels are kept hidden during model induction). We wish to learn a responder-prediction model from existing aggregate-label ALLEGRO data and unlabeled CONCERTO data and use the model to identify likely laquinimod-responders for the CONCERTO trial. The distribution of responders and non-responders revealed in Figure 9 suggests this problem would be daunting even if individual-level labels were available from the ALLEGRO trial.

**Figure 9.**
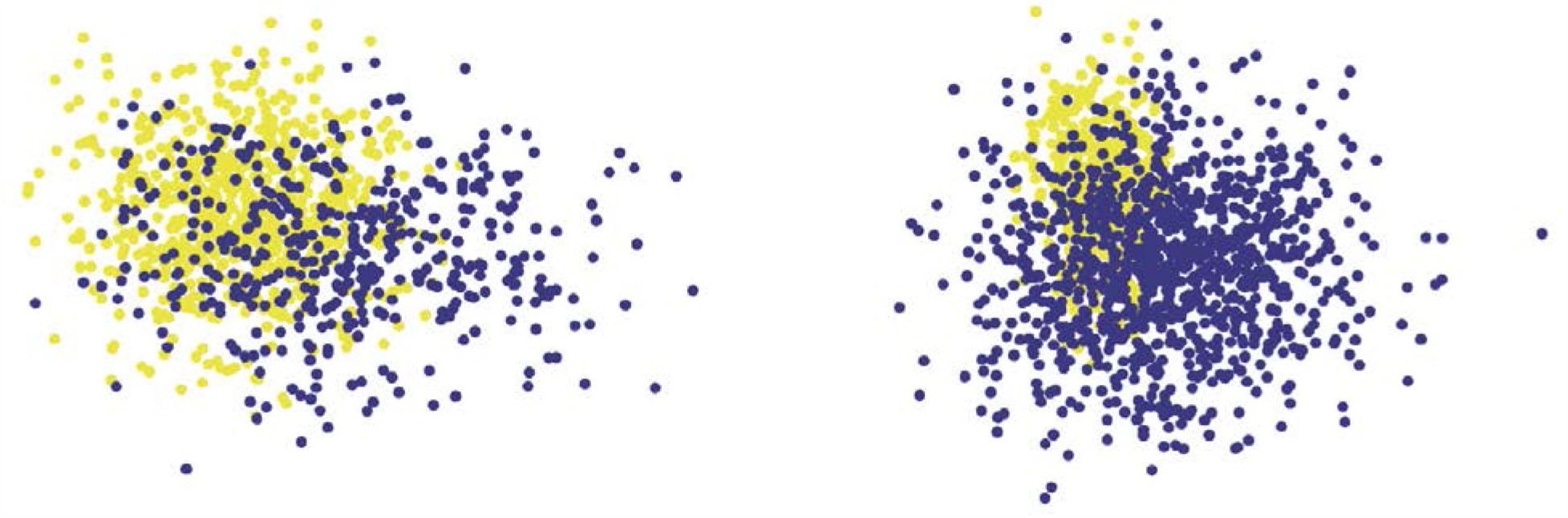
Responders and non-responders for the MS DMT laquinimod. Plots are two-dimensional feature-space projections of the ground-truth responders (yellow) and non-responders (blue) for the ALLEGRO trial (left, 1101 participants) and CONCERTO trial (right, 1456 participants).

**Figure 10.**
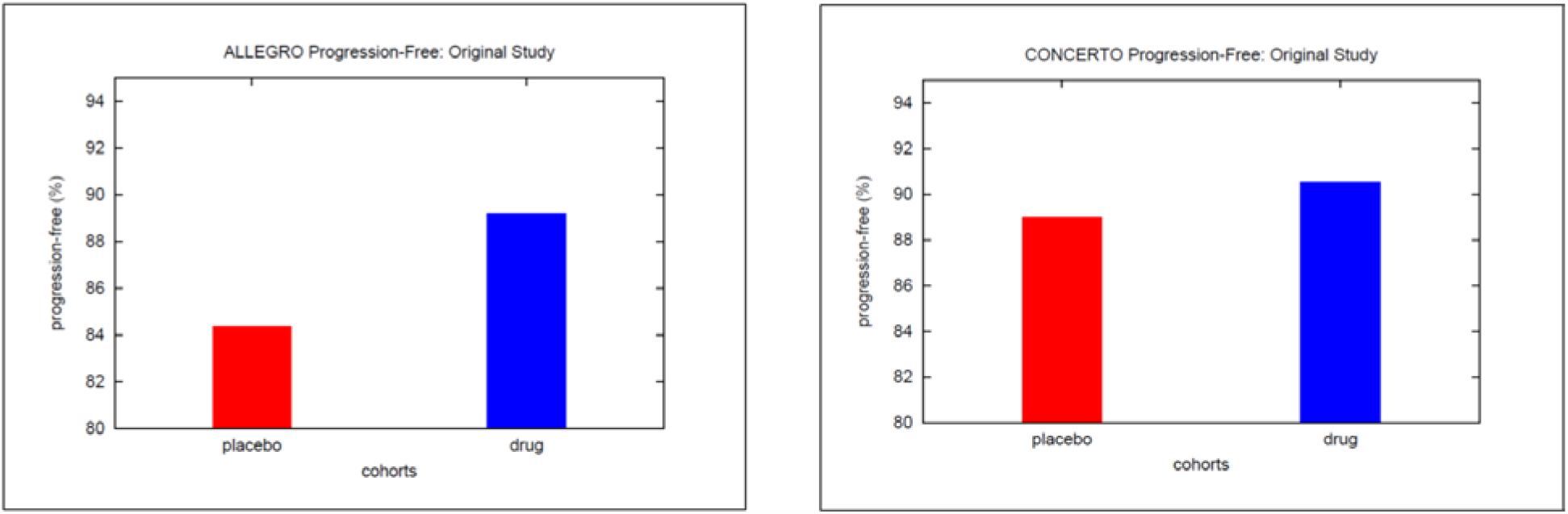
Responders and non-responders in the (original) laquinimod/placebo clinical trial arms. Plots display fractions of disability-progression-free patients in the placebo (red) and laquinimod (blue) cohorts for the ALLEGRO (left) and CONCERTO (right) trials.

Figure 10 shows the fraction of disability-progression-free patients (responders) in the laquinimod (blue) and placebo (red) cohorts for the ALLEGRO trial (left) and CONCERTO trial (right). Observe that with ALLEGRO there is a significantly larger fraction of progression-free patients in the laquinimod-treated arm, and that the difference in responder-fractions between laquinimod and placebo arms is smaller (and not significant) for CONCERTO. This discussion motivates the question pursued in this experiment: is it feasible to predict, before trial launch, which CON-CERTO participants will be laquinimod responders and leverage these predictions to demonstrate treatment efficacy for a subpopulation of trial patients?

The main results obtained in the experiments are presented in Figure 11. The left two bars of the plot show trial outcomes for those CONCERTO patients predicted to be non-responders by Algorithm LIPAL (trained with aggregate-label ALLEGRO data), and the right two bars report outcomes for the predicted responders. More precisely, the left bars display fractions of disability progression-free patients among predicted non-responders who had been placed into the placebo (red) and laquinimod (blue) trial arms, and the right two bars show the same quantities for predicted responders. It is seen from Figure 11 that, while there is no (statistical) difference in treatment effects for predicted non-responders, there is significant benefit to laquinimod treatment for predicted responders. These results support the feasibility of identifying likely DMT-responders in advance and using these predictions for precision clinical trials.

**Figure 11.**
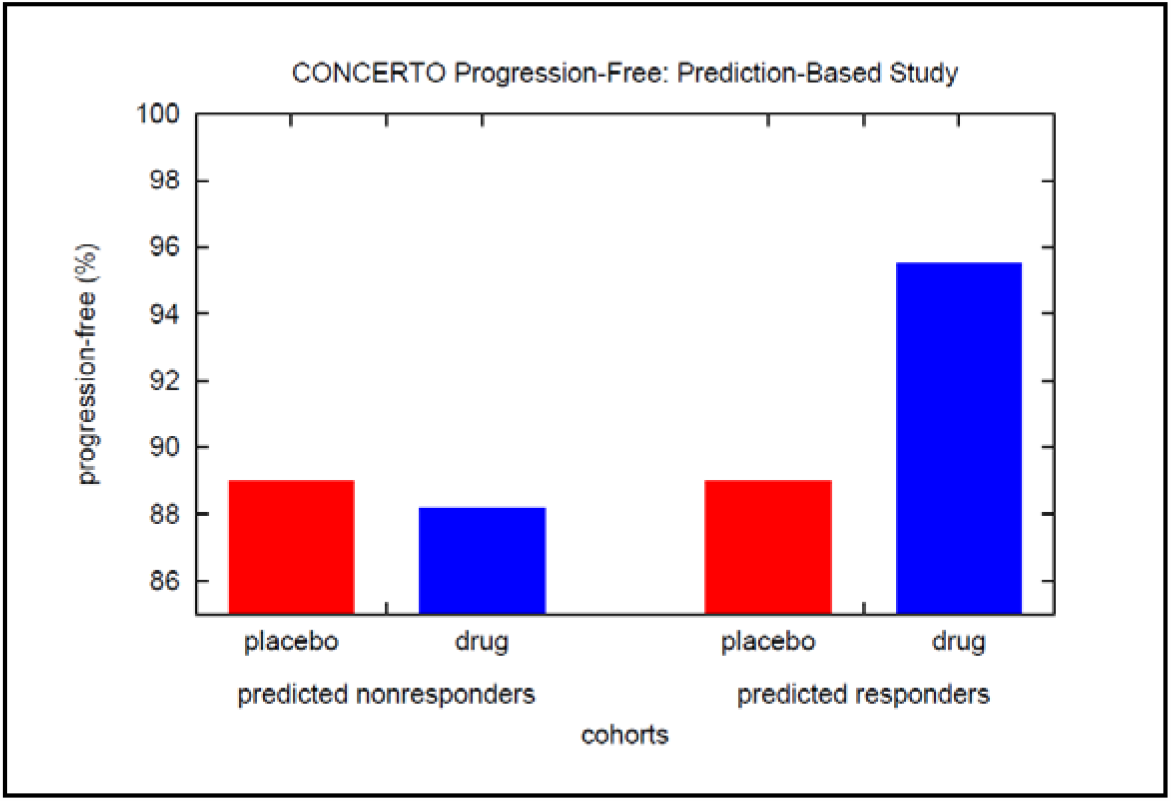
Behavior of predicted responders/non-responders in CONCERTO trial. Right two bars show the fractions of progression-free patients among predicted responders who had been placed into the placebo (red) and laquinimod (blue) trial arms. Left two bars depict the same quantities for the predicted non-responders.

Knowing which features captured in clinical trials data can be used to detect DMT-responders is of value in various applications. We estimate feature predictive power using forward/backward-stepwise analyses [7] and find that predictive features include demographics attributes (age, gender), number of MS relapses in the previous year, baseline brain volume, and baseline MRI lesion activity (gadolinium-enhancement). Note that all these features can be measured in the course of routine care.

## 6. Concluding Remarks

This paper presents a new machine learning algorithm that enables *individual-level* prediction models to be induced from *aggregate-level* labels, which are abundant in many health domains. The utility of the proposed learning methodology for personalized medicine applications is demonstrated by: i.) leveraging geographically-aggregated mental health statistics to build a screening tool which detects individuals suffering from depression based upon their Twitter activity; ii.) designing decision-support systems that exploits aggregate clinical trials data on MS drugs to predict optimal individualized therapy and find those patients likely to benefit from an experimental therapy.

Future work will include exploring the extent to which the approach can be deployed to accomplish additional precision medicine tasks and combining Algorithm LIPAL with other, complementary lightly-supervised learning strategies [21,23,26,36].

## Data Availability

The data used in the case studies in the submitted article are available through the referenced works.

## Notes

### Competing Interest Statement

Kristin Glass and Rich Colbaugh are shareholders in Volv Global.

### Funding Statement

Volv Global supported the research described in the submitted article.

### Author Declarations

The research reported in the submission is a retrospective study of publically available information.

